# The Unequal Burden of the Covid-19 Pandemic: Racial/Ethnic Disparities in US Cause-Specific Mortality

**DOI:** 10.1101/2021.08.25.21262636

**Authors:** Anneliese N. Luck, Samuel H. Preston, Irma T. Elo, Andrew C. Stokes

## Abstract

**Objectives:** To quantify changes in all-cause and cause-specific mortality by race and ethnicity between 2019 and 2020.

**Methods:** Using 2019 and 2020 provisional death counts from the National Center for Health Statistics and population estimates from the US Census Bureau, we estimate age-standardized death rates by race/ethnicity and attribute changes in mortality to various causes of death. We also examine how patterns of change across racial/ethnic groups vary by age and sex.

**Results:** Covid-19 death rates in 2020 were highest in the Hispanic community whereas Black individuals had the largest increase in all-cause mortality between 2019 and 2020. Increases in mortality from heart disease, diabetes, and external causes of death accounted for the adverse trend in all-cause mortality within the Black population. Percentage increases in all-cause mortality were similar for men and women and for ages 25-64 and 65+ for Black and White populations, but increases were greatest for working-aged men among the Hispanic population.

**Conclusions:** Examining increases in non-Covid-19 causes of death is essential for fully capturing both the direct and indirect impact of the Covid-19 pandemic on racial/ethnic mortality disparities.

## INTRODUCTION

The onset of the Covid-19 pandemic in 2020 led to a sharp rise in mortality in the United States, with Black and Hispanic communities absorbing a disproportionate share of the mortality impact.^1–3^ Recent research suggests that the 2020 life expectancy drop eliminated the progress made in the past decade in narrowing the Black-White life expectancy gap and nearly erased the Hispanic life expectancy advantage.^2,3^

Several factors may place Black and Hispanic populations more at risk of Covid-19 infection and mortality. These factors include disproportionate representation in frontline jobs with minimal workplace protections, a higher prevalence of comorbid conditions, and unequal access to health care^3–5^ – all of which reflect a system of structural racism that has long shaped unequal treatment and access to health equity in the United States.^6,7^

Although the disparities in Covid-19 deaths are well-documented, studies that only consider deaths directly attributable to Covid-19 may obscure the overall magnitude of racial/ethnic disparities in COVID-19 related mortality for several reasons. First, Covid-19 death counts do not capture the indirect impact of the pandemic on mortality, driven by increased economic and housing hardship, worsened mental health, and reductions in healthcare access.^8,9^ Second, official Covid-19 death counts fail to capture Covid-19 deaths that were misclassified to other causes of death. Such deaths inappropriately ascribed to other causes may be especially prevalent among racial/ethnic groups who have historically received less adequate medical attention.^10^

This study investigates the total mortality impact of the Covid-19 pandemic on racial /ethnic mortality disparities. Using age-standardized death rates for Non-Hispanic White, Non-Hispanic Black, and Hispanic individuals (hereafter referred to as White, Black, and Hispanic), we examine patterns of mortality change between 2019 and 2020 by underlying cause of death, age, and sex and identify the causes of death with the greatest impact on changes in racial/ethnic mortality differentials.

## METHODS

All analyses were conducted using a combination of publicly available datasets from the National Center for Health Statistics (NCHS) and the United States Census Bureau. Provisional monthly death counts for January-December 2019 and 2020, were obtained for the US population aged 25+ by available underlying causes of death, 10-year age groups (25-34, 35-44, … 75-85, 85+), sex, and single-race coded categories for race/ethnicity.^11^ Covid-19 deaths were restricted to deaths in which Covid-19 was assigned as the underlying cause. This approach is the same as in Ahmad & Anderson (2021) and in Glei (2021).^12,13^ The inclusion of Covid-19 deaths not assigned as an underlying cause would have raised 2020 Covid-19 deaths by 9.6% and would have prevented us from using a mutually exclusive and exhaustive set of 13 underlying causes, one of which was Covid-19. We also created a residual all other cause-of-death category. Mid-year (July 1^st^) population estimates by age, sex, and race/ethnicity were obtained from the Census Bureau’s monthly national-level population estimates.^14^

Crude death rates were age-standardized using the mid-year 2020 national age distribution for population aged 25+ as well as for ages 25-64 and 65+. All-cause and cause-specific age-standardized death rates (ASDR) were derived by sex and by race/ethnicity for White, Black, and Hispanic populations. We start at age 25 because the number of deaths from Covid-19 in 2020 below age 25 was relatively small. The number of all-cause deaths below age 25 in 2020 was 2% below the average for 2015-19.^15^ We use age-standardized rates to avoid spurious changes in numbers of deaths or crude death rates resulting from population growth and/or aging.^16^

All data aligned with the NCHS’s single-race coding schema for racial/ethnic categories in the provisional mortality file, where Hispanic corresponds to all persons listed as Hispanic regardless of race, but non-Hispanic White and non-Hispanic Black correspond to those who selected White and Black *only*, respectively.

We assume that the difference in age-standardized all-cause mortality rates between 2019 and 2020 reflects the effect of the Covid-19 pandemic on mortality, aligning with previous descriptive work on excess mortality in 2020.^13^ Alternative approaches have compared 2020 mortality to mortality projected in 2020 on the basis of mortality rates in years prior to 2020.^17,18^ The absence of single-race death counts before 2018 prevents us from using the time-series approach on single-race data.^19^ Instead, we used bridged-race deaths to investigate how results would change if, instead of using 2019 rates for comparison to 2020 rates, we had used predicted 2020 rates based on a time series fitted to 2015-2019 observations. This analysis investigates the sensitivity of the results for all-cause mortality and for selected causes of death whose changes accounted for largest changes in all-cause mortality between 2019 and 2020 [Appendix Table F].

## RESULTS

For the entire United States, the number of deaths at ages 25+ rose by 19.0% between 2019 and 2020 [Appendix Table A] and the age-standardized death rate at ages 25+ rose by 17.0%. The age-standardized percentage varied sharply among racial/ethnic groups, increasing by only 13.0% among the White population compared to 27.2% and 39.3% among the Black and Hispanic population, respectively.

The ordering of the mortality change by racial/ethnic group depends on whether we focus on the absolute or the percentage change in the age-standardized death rate. While Hispanic individuals suffered the largest percentage increase in age-standardized mortality, the ranking of Hispanic and Black populations reverses in relation to absolute increases [Table 1]. We primarily feature the absolute change in this paper because it is the most direct measure of the change in the frequency of death per person.

**Table 1.**
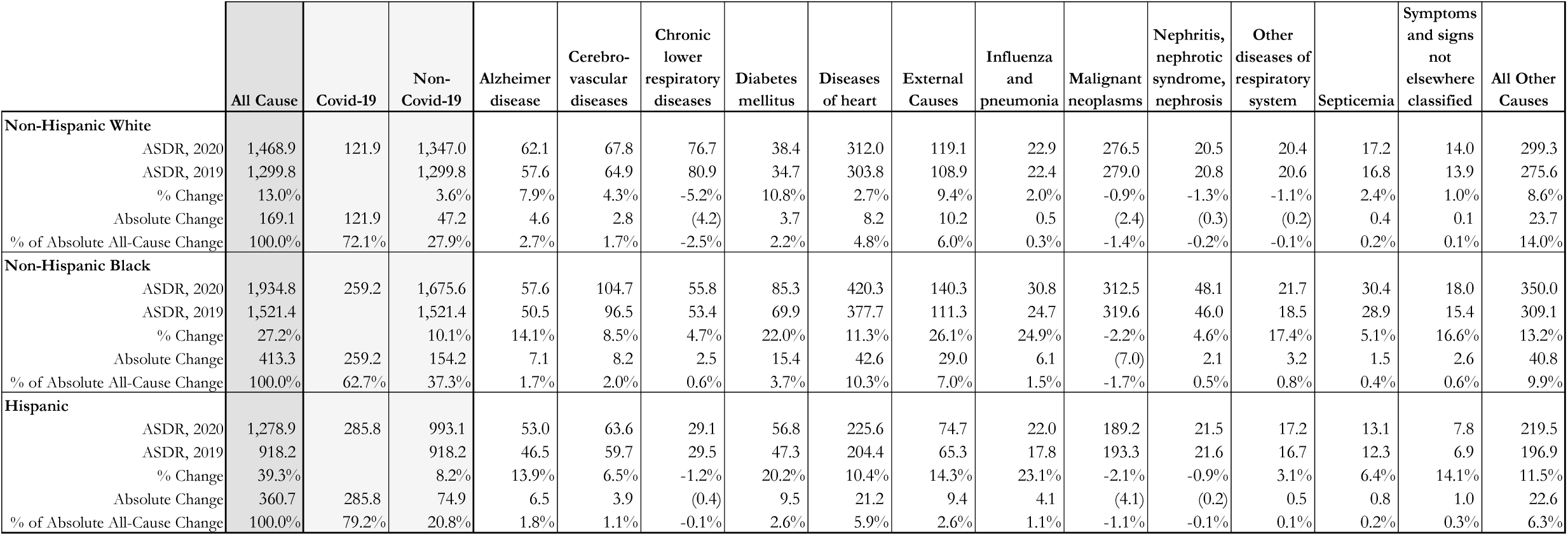
Change in ASDR (ages 25+) by Cause of Death and Race/Ethnicity, Both Sexes.

|  | All Cause | Covid-19 | Non-Covid-19 | Alzheimer disease | Cerebro-vascular diseases | Chronic lower respiratory diseases | Diabetes mellitus | Diseases of heart | External Causes | Influenza and pneumonia | Malignant neoplasms | Nephritis, nephrotic syndrome, nephrosis | Other diseases of respiratory system | Septicemia | Symptoms and signs not elsewhere classified | All Other Causes |
| --- | --- | --- | --- | --- | --- | --- | --- | --- | --- | --- | --- | --- | --- | --- | --- | --- |
| **Non-Hispanic White** |  |  |  |  |  |  |  |  |  |  |  |  |  |  |  |  |
| ASDR, 2020 | 1,468.9 | 121.9 | 1,347.0 | 62.1 | 67.8 | 76.7 | 38.4 | 312.0 | 119.1 | 22.9 | 276.5 | 20.5 | 20.4 | 17.2 | 14.0 | 299.3 |
| ASDR, 2019 | 1,299.8 |  | 1,299.8 | 57.6 | 64.9 | 80.9 | 34.7 | 303.8 | 108.9 | 22.4 | 279.0 | 20.8 | 20.6 | 16.8 | 13.9 | 275.6 |
| % Change | 13.0% |  | 3.6% | 7.9% | 4.3% | -5.2% | 10.8% | 2.7% | 9.4% | 2.0% | -0.9% | -1.3% | -1.1% | 2.4% | 1.0% | 8.6% |
| Absolute Change | 169.1 | 121.9 | 47.2 | 4.6 | 2.8 | (4.2) | 3.7 | 8.2 | 10.2 | 0.5 | (2.4) | (0.3) | (0.2) | 0.4 | 0.1 | 23.7 |
| % of Absolute All-Cause Change | 100.0% | 72.1% | 27.9% | 2.7% | 1.7% | -2.5% | 2.2% | 4.8% | 6.0% | 0.3% | -1.4% | -0.2% | -0.1% | 0.2% | 0.1% | 14.0% |
| **Non-Hispanic Black** |  |  |  |  |  |  |  |  |  |  |  |  |  |  |  |  |
| ASDR, 2020 | 1,934.8 | 259.2 | 1,675.6 | 57.6 | 104.7 | 55.8 | 85.3 | 420.3 | 140.3 | 30.8 | 312.5 | 48.1 | 21.7 | 30.4 | 18.0 | 350.0 |
| ASDR, 2019 | 1,521.4 |  | 1,521.4 | 50.5 | 96.5 | 53.4 | 69.9 | 377.7 | 111.3 | 24.7 | 319.6 | 46.0 | 18.5 | 28.9 | 15.4 | 309.1 |
| % Change | 27.2% |  | 10.1% | 14.1% | 8.5% | 4.7% | 22.0% | 11.3% | 26.1% | 24.9% | -2.2% | 4.6% | 17.4% | 5.1% | 16.6% | 13.2% |
| Absolute Change | 413.3 | 259.2 | 154.2 | 7.1 | 8.2 | 2.5 | 15.4 | 42.6 | 29.0 | 6.1 | (7.0) | 2.1 | 3.2 | 1.5 | 2.6 | 40.8 |
| % of Absolute All-Cause Change | 100.0% | 62.7% | 37.3% | 1.7% | 2.0% | 0.6% | 3.7% | 10.3% | 7.0% | 1.5% | -1.7% | 0.5% | 0.8% | 0.4% | 0.6% | 9.9% |
| **Hispanic** |  |  |  |  |  |  |  |  |  |  |  |  |  |  |  |  |
| ASDR, 2020 | 1,278.9 | 285.8 | 993.1 | 53.0 | 63.6 | 29.1 | 56.8 | 225.6 | 74.7 | 22.0 | 189.2 | 21.5 | 17.2 | 13.1 | 7.8 | 219.5 |
| ASDR, 2019 | 918.2 |  | 918.2 | 46.5 | 59.7 | 29.5 | 47.3 | 204.4 | 65.3 | 17.8 | 193.3 | 21.6 | 16.7 | 12.3 | 6.9 | 196.9 |
| % Change | 39.3% |  | 8.2% | 13.9% | 6.5% | -1.2% | 20.2% | 10.4% | 14.3% | 23.1% | -2.1% | -0.9% | 3.1% | 6.4% | 14.1% | 11.5% |
| Absolute Change | 360.7 | 285.8 | 74.9 | 6.5 | 3.9 | (0.4) | 9.5 | 21.2 | 9.4 | 4.1 | (4.1) | (0.2) | 0.5 | 0.8 | 1.0 | 22.6 |
| % of Absolute All-Cause Change | 100.0% | 79.2% | 20.8% | 1.8% | 1.1% | -0.1% | 2.6% | 5.9% | 2.6% | 1.1% | -1.1% | -0.1% | 0.1% | 0.2% | 0.3% | 6.3% |

Figure 1 illustrates the contribution of Covid-19 and causes *other than* Covid-19 to changes in all-cause mortality between 2019 and 2020. Although the Black population had the largest increase, the reason was not that Black individuals had the highest Covid-19 death rates. The Hispanic population aged 25+ saw the highest Covid-19 death rate in 2020 (286/100,000), compared to 259/100,000 for the Black population and 122/100,000 for the White population [Table 1].

**Figure 1:**
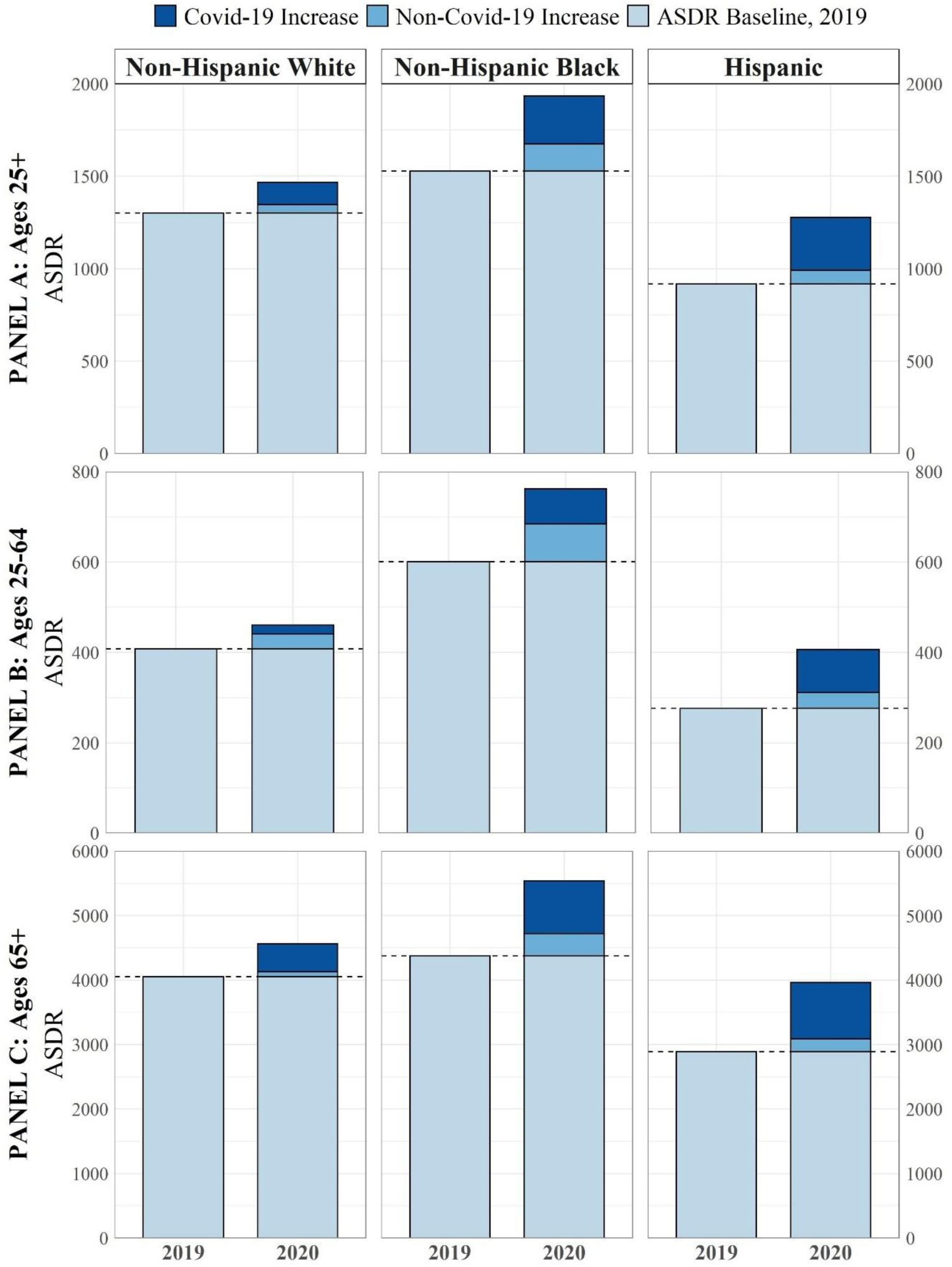
Age-Standardized Death Rates (ASDR) by Race/Ethnicity, 2019-2020. Note: Figure shows All-Cause ASDR by race/ethnicity (both sexes) in 2019 and 2020, with Covid-19 and non-Covid-19 increases in 2020 highlighted. Y-axis scales change between age group panels.

The figure also reveals that mortality from underlying causes of death other than Covid-19 rose for all three groups. The Black population aged 25+ saw the largest increase in mortality from causes other than Covid-19, with an increase of 154/100,000, followed by the Hispanic population (75/100,000) and the White population (47/100,000). Of the total increase in mortality between 2019 and 2020, Covid-19 deaths accounted for 62.7% of the increase among Black, 72.1% among White, and 79.2% among Hispanic individuals.

Other causes of death contributing to the mortality increase between 2019 and 2020 are shown in Table 1 and in Figure 2. Apart from Covid-19 and the residual category, the three largest increases in mortality for Black and Hispanic individuals were from heart disease, diabetes, and external causes of death. These were three of the four largest increases among White populations as well, along with Alzheimer’s disease. But the magnitude of the increases was markedly different; Black individuals experienced the greatest increases from all three causes of death, with ratios of increase ranging from 1.6 to 5.2. In fact, for *all* causes of death except malignant neoplasms, where mortality fell, Black populations suffered from larger mortality increases than White or Hispanic populations.

**Figure 2:**
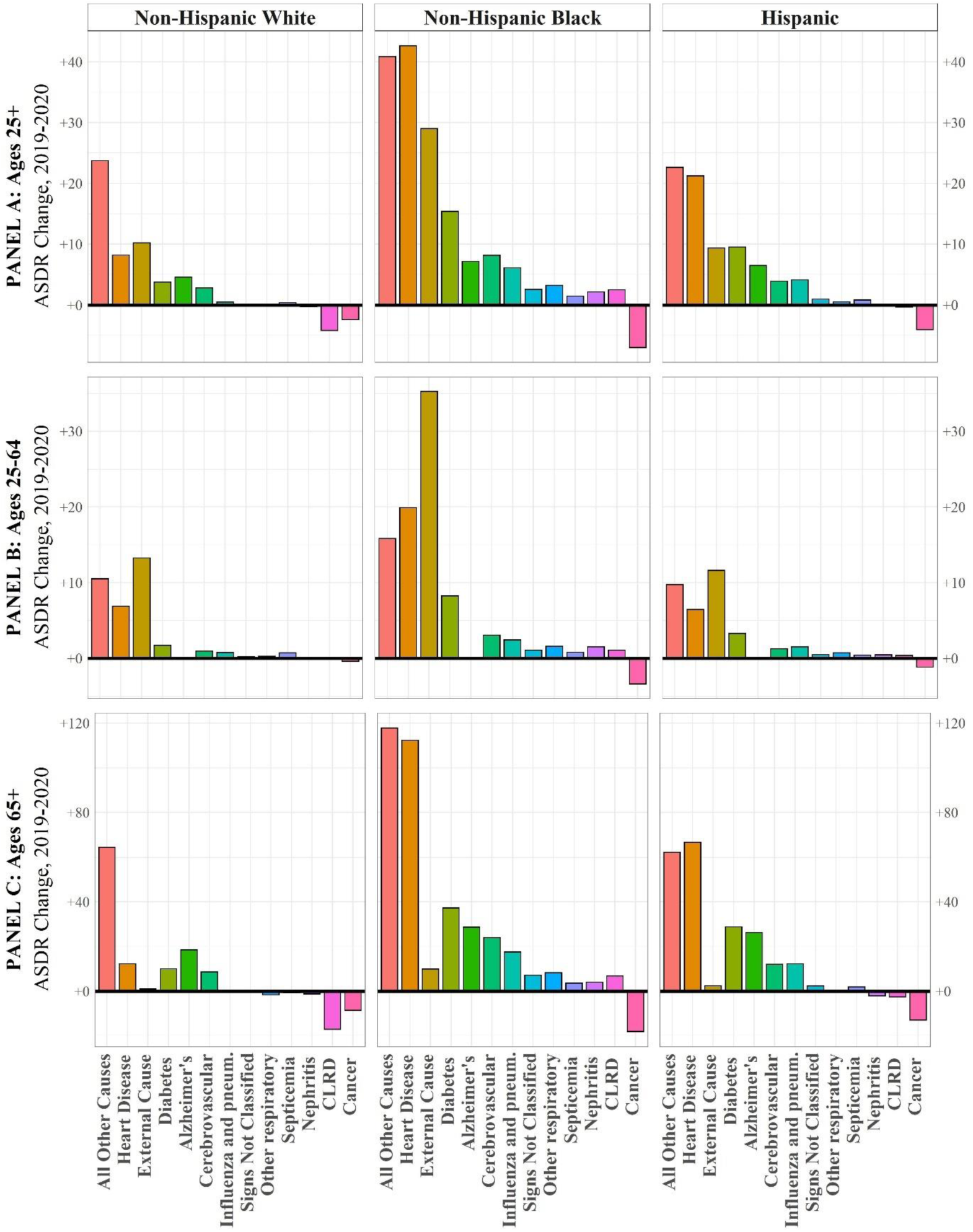
Change in Causes of Death other than Covid-19 by Race/Ethnicity, 2019-2020. Note: Figure shows absolute ASDR change by race/ethnicity (both sexes) for all non-Covid-19 causes of death in 2020, relative to 2019. Y-axis scales change between age group panels.

The same four cause-of-death categories dominate racial/ethnic mortality disparities [Appendix Table B]. Fifty-seven percent of the increase in age-standardized Black mortality relative to White mortality was attributable to Covid-19, while 14.1% was attributable to heart disease, 7.7% to external causes, and 4.8% to diabetes. The change in the Hispanic and White mortality gap was overwhelmingly attributable to Covid-19, which accounted for 85.5% of the relative rise in Hispanic mortality. Greater increases for Black mortality relative to Hispanic mortality from heart disease, external causes, and diabetes more than offset the greater increase from Covid-19 in the Hispanic population.

Age-specific changes in mortality are also patterned by race/ethnicity [Appendix Table C]. For White and Black individuals, mortality rose by the same percentage at working ages (25-64) and older ages (65+), 13% and 27%, respectively. These age-patterns of mortality change reflect pre-existing age-patterns in 2019. Among the Hispanic population, however, the increase in the death rate at working ages, 47%, was substantially greater than the increase at ages 65+, 37%. Working-age Hispanic individuals, ages 25-64, had by far the sharpest percentage increase in mortality of any age/race/ethnicity group.

Since the age-pattern of mortality from Covid-19, like that of all causes of death combined, is skewed towards older individuals, mortality changes for ages 25+ closely reflect those that occurred at ages 65+. The story at ages 25-64 is quite different. Although Covid-19 accounted for most of the all-cause mortality increase for the Hispanic population (72.9%), it was responsible for only 36.5% of the increase among White individuals and 46.9% among Black individuals. For all three race/ethnic groups, the cause of death contributing the most to increases in mortality between 2019 and 2020 at ages 25-64, apart from Covid-19, was external causes of death. The rise in external-cause mortality for the Black population was 2.7-3.0 times the increase for the other two groups. Increases in mortality from heart disease and diabetes were the third and fourth largest contributors to all-cause mortality increases at age 25-64 for all three groups.

The pattern of change in mortality between 2019 and 2020 was relatively similar for men and women [Appendix Tables D and E]. Hispanic individuals showed the greatest disparity between men (44.0% increase) and women (33.4% increase), a difference that was exaggerated at working-ages (51.9% vs. 37.9%). Sex differences in the contribution of Covid-19 to mortality change were also minor.

One cause of death showed a sharp differentiation between the sexes. For all three groups, mortality from external causes increased much more in 2020 for men than for women. This distinction was especially pronounced in the working ages, where the rise in mortality was 19.4/100,00 for males vs. 3.5 for females among Hispanic, 19.2 vs. 7.2 among White, and 57.1 vs. 15.0 among Black populations.

Although our analysis has focused on absolute changes in death rates, percentage changes depicted in Figure 3 shed additional light. The Figure presents scatter diagrams of percentage changes in mortality at ages 25+ for different causes of death for White individuals on the x-axis and either Black or Hispanic individuals on the y-axis. Points above the 45-degree line indicate causes of death for which Black or Hispanic populations had a higher percentage increase (or smaller decline) than their White peers. Nearly all points are above the 45-degree lines. For both Black and Hispanic communities, the highest percentage increase relative to White communities pertains to influenza and pneumonia, a category with symptoms similar to those of Covid-19. Increases in mortality from diabetes, heart disease, Alzheimer’s, chronic lower respiratory diseases, external causes, other respiratory diseases, and abnormal signs and symptoms are also higher for both Hispanic and Black populations as compared to the White population [Table 1].

**Figure 3:**
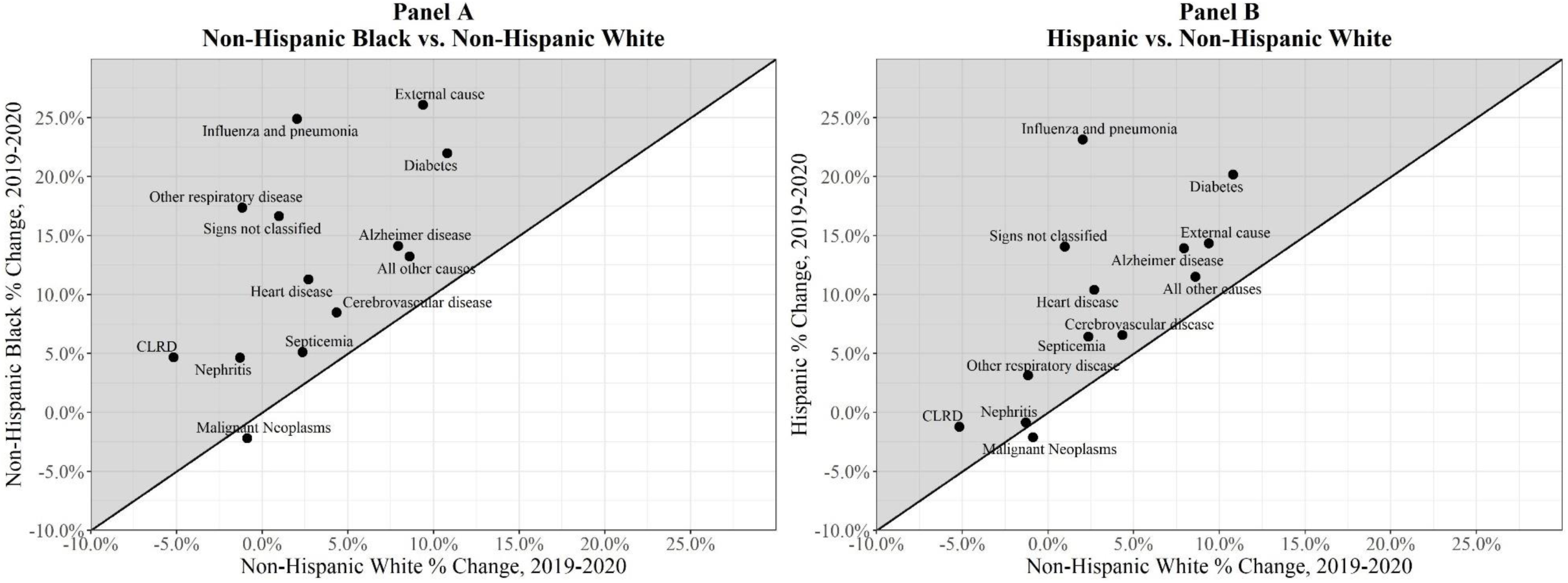
Racial/Ethnic Comparisons of Non-Covid-19 ASDR Change (Both Sexes, Ages 25+), 2019-2020. Note: Figure shows racial/ethnic comparisons ASDR percent change for all non-Covid-19 causes of death in 2020, relative to 2019. Shaded area indicates where percent increase was greater for Black (Panel A) and Hispanic (Panel B) than their White peers.

## DISCUSSION

Although mortality rose sharply for White, Black, and Hispanic populations between 2019 and 2020, the experience of that increase varied greatly by race/ethnicity. The largest all-cause mortality increase was recorded for the Black population, but the reason was not the number of deaths assigned to Covid-19 as an underlying cause of death. Hispanic populations, in fact, experienced the largest increase in Covid-19 mortality. Instead, Black individuals had by far the largest increase in death rates from other causes of death, nearly double that among Hispanic individuals and more than triple that among White individuals.

### High Covid-19 Mortality among Hispanic Communities

The Hispanic Covid-19 death rate in 2020 was around 1.1 times higher than that of their Black peers and 2.3 times higher than their White peers [Table 1]. These findings are consistent with other evidence that has emerged surrounding the particularly adverse impact of the pandemic on Hispanic mortality.^20–22^

The high mortality of Hispanic individuals from Covid-19 appears to be primarily attributable to a high incidence of infection and secondarily to a high case-fatality rate. A study of Covid-19 incidence among 50 million health system enrollees revealed a per capita infection rate of 143/1000,00 for Hispanic individuals, relative to 107 and 46 among their Black and White peers, respectively.^23^ The relative rate ratios of infection are even higher than the respective ratios of Covid-19 mortality [Appendix Table B], suggesting that infection rates are driving mortality differentials.^24^

A large body of research has shown how structural and systemic racism routinely disadvantages communities of color,^6,7^ shaping a variety of institutional factors that place Hispanic populations at heightened risk of Covid-19 incidence. The Hispanic community is disproportionately represented in essential occupations with high-exposure to Covid-19 and limited workplace protections.^21,22^ Additionally, Hispanic individuals are more likely to live in denser, multigenerational households, magnifying the effect of occupational exposure.^5,20^ These factors may be particularly influential within the immigrant community, where an even more pronounced mortality disadvantage during the pandemic has been found.^20^ We show that all-cause and Covid-19 mortality was particularly inflated among the Hispanic working-aged and male populations, underscoring the potential saliency of workplace conditions in producing a distinct Covid-19 mortality disadvantage for the Hispanic community.

Reduced access to adequate health care also likely contributes towards higher Covid-19 case-fatality risks among Hispanic individuals. Hispanic communities have the lowest rate of health insurance coverage of any racial/ethnic group in the United States, with nearly 20% uninsured, compared to just 5% of the White population.^5^ Additionally, language barriers, distrust and fear of healthcare institutions driven by broader anti-immigrant rhetoric and policy, and underlying financial hardship faced by Hispanic individuals, particularly those in the immigrant community, have all been linked to worse healthcare access and quality, potentially amplifying the impact of Covid-19 on mortality outcomes.^5^ These health care barriers, coupled with a higher prevalence of comorbid conditions, such as diabetes and heart disease, may worsen Covid-19 outcomes once infection occurs.^22^

### High Mortality from Non-Covid-19 Causes of Death Among Black Individuals

Despite having levels of Covid-19 incidence and mortality below those of Hispanic individuals, Black individuals saw the largest absolute increase in all-cause mortality, a pattern consistent with previous work.^25,26^ This increase is attributable to causes other than Covid-19, highlighting the ways that the pandemic has indirectly shaped mortality among Black communities during the Covid-19 pandemic.

Increases in heart disease and diabetes for Black individuals were more than four times those of their White peers and 1.6-2.0 times those of their Hispanic peers [Table 1]. One explanation offered for these trends is that the pandemic disrupted access to personal support networks and health care in ways that hindered disease management for people with these chronic diseases.^12,13^

Fear of infection, loss of health insurance, reductions in visits to physicians and in elective procedures have been noted as indirect effects of the pandemic.^9^ These factors are more salient among Black individuals for whom the pre-pandemic prevalence of serious heart disease, hypertension and diabetes exceeded that among White individuals.^27^

Mortality from external causes of death also rose sharply among Black individuals. This increase was nearly three times greater for the Black population than for the White or Hispanic population, a distinction most pronounced among working-age males. Unfortunately, the data do not currently permit us to distinguish among external causes. Previous research has shown that the principal increase in mortality within this category between 2019 and 2020 pertains to drug-related mortality,^12,13^ and there are indications that this rise may be more pronounced among Black and Hispanic communities.^28^ There is also emerging suggestive evidence of suicide increases particularly among Black and Hispanic communities.^29^ Growth in external-cause mortality draws attention to the ways in which social isolation, economic instability, and treatment disruption may have exacerbated “deaths of despair” among working-age Americans,^30,31^ warranting particular concern for Black and Hispanic communities who have absorbed a disproportionate share of these forms of disadvantage during the pandemic.

### Possible Errors in Diagnosis and Coding of Underlying Cause of Death

Another explanation of changing death rates in 2020 relates to the process by which underlying cause of death is assigned on death certificates. Some jurisdictions across the U.S., especially early in the pandemic, required a positive test result for a death to be assigned to COVID-19 and thus an absence of testing may have led to a death being assigned to a comorbid chronic condition such as diabetes or heart disease.^32^ Accurate cause of death assignment is also likely complicated by the large number of home deaths, especially among racial/ethnic minority populations.^33^ Deaths occurring at home are more likely to be certified by a local coroner, who can lack medical training and has limited resources for performing post-mortem testing or autopsies.^34^

Influenza/pneumonia represents another cause of death for which deaths may be spuriously inflated by the Covid-19 pandemic. Early in the pandemic, many deaths from Covid-19 were assigned to influenza/pneumonia.^17^ As suggested previously in Figure 3, such mis-assignment was likely to be much more common among Black and Hispanic than White individuals.

### Limitations

Our analysis had several limitations. First, deaths included in this analysis were available from the NCHS through August 3, 2021 and it is possible that delays in registration will result in additional deaths being reported for 2020. Such delays are especially likely for external causes of death for which certification often involves multiple authorities. Second, the present paper was limited to Black, Hispanic, and White adults, as data quality was greatest in these groups and the populations were sufficiently large to generate stable mortality rates by age, sex, and cause of death. Future research should incorporate analyses of American Indian and Alaskan Natives and other racial/ethnic groups that experienced significant adverse mortality trends during the Covid-19 pandemic.^25^

A third limitation involves uncertainty about how deaths attributable to the Covid-19 should be estimated. Alternative methods of estimating what mortality would have been in 2020 in the absence of the Covid-19 pandemic will produce different estimates of the pandemic’s impact. The present analysis employed direct comparisons of mortality between 2019 and 2020 in order to maintain consistency in single-race coding across years.^13^ We present evidence in the Appendix that 2019 death rates at ages 25+ and 65+ from all-causes, heart disease and diabetes are very consistent with predicted 2020 mortality levels using a linear modeling approach based on bridged-race deaths between 2015 and 2019. In contrast, estimates of changes in mortality from external causes are sensitive to whether prior mortality trends are incorporated into estimates of expected deaths in 2020. This sensitivity is likely the result from the rising drug overdose mortality already evident between 2015 and 2019, a trend that may have continued into 2020.^12^

Finally, as noted earlier, our study is based on data in which Covid-19 is listed as the underlying cause of death and detailed cause of death information available in the publicly released NCHS data is limited to 13 select causes of death.^35^ Thus our residual cause-of-death category comprised a small but not an insignificant portion of the increase in all-cause mortality at ages 25+(6.3% for Hispanic, 9.7% for Black, and 13.9% for White populations). Future research should further explore how varying time-series approaches and more detailed cause-of death information may affect the estimates presented here.

In conclusion, our findings suggest that the overall effect of the Covid-19 pandemic on racial/ethnic disparities was much larger than that captured by official Covid-19 death tallies alone. Black individuals had exceptionally large increases in non-Covid-19 causes of death during the pandemic, including from heart disease, diabetes, and external causes of death, and assessment of these deaths is critical to a full accounting of the pandemic’s disparate impacts on population health.

## Data Availability

All data is publicly available through the National Center for Health Statistics and US Census Bureau.

https://data.cdc.gov/NCHS/AH-Monthly-Provisional-Counts-of-Deaths-for-Select/65mz-jvh5

https://wonder.cdc.gov/ucd-icd10.html

https://www.census.gov/data/tables/time-series/demo/popest/2010s-national-detail.html

## SUPPLEMENTARY APPENDIX

**Table A.** All-Cause Deaths, Population, and ASDR for ages 25+ by Race/Ethnicity and Sex.

|  | Deaths |  |  | Population |  |  | ASDR |  |  |
| --- | --- | --- | --- | --- | --- | --- | --- | --- | --- |
|  | Male | Female | Total | Male | Female | Total | Male | Female | Total |
| **Total Population** |  |  |  |  |  |  |  |  |  |
| 2020 | 1,731,439 | 1,593,707 | 3,325,146 | 109,725,881 | 116,909,132 | 226,635,013 | 1,747 | 1,230 | 1,467 |
| 2019 | 1,435,071 | 1,359,755 | 2,794,826 | 108,906,904 | 116,099,507 | 225,006,411 | 1,479 | 1,064 | 1,254 |
| Absolute Change | 296,368 | 233,952 | 530,320 | 818,977 | 809,625 | 1,628,602 | 267 | 166 | 213 |
| Ratio | 1.207 | 1.172 | 1.190 | 1.008 | 1.007 | 1.007 | 1.181 | 1.156 | 1.170 |
| **Non-Hispanic White** |  |  |  |  |  |  |  |  |  |
| 2020 | 1,262,029 | 1,196,340 | 2,458,369 | 70,348,355 | 74,383,706 | 144,732,061 | 1,727 | 1,245 | 1,469 |
| 2019 | 1,098,199 | 1,058,330 | 2,156,529 | 70,293,191 | 74,355,654 | 144,648,845 | 1,521 | 1,109 | 1,300 |
| Absolute Change | 163,830 | 138,010 | 301,840 | 55,164 | 28,052 | 83,216 | 206 | 136 | 169 |
| Ratio | 1.149 | 1.130 | 1.140 | 1.001 | 1.000 | 1.001 | 1.135 | 1.123 | 1.130 |
| **Non-Hispanic Black** |  |  |  |  |  |  |  |  |  |
| 2020 | 226,329 | 206,040 | 432,369 | 12,612,934 | 14,600,484 | 27,213,418 | 2,411 | 1,581 | 1,935 |
| 2019 | 171,304 | 159,967 | 331,271 | 12,452,724 | 14,436,702 | 26,889,426 | 1,877 | 1,258 | 1,521 |
| Absolute Change | 55,025 | 46,073 | 101,098 | 160,210 | 163,782 | 323,992 | 534 | 323 | 413 |
| Ratio | 1.321 | 1.288 | 1.305 | 1.013 | 1.011 | 1.012 | 1.284 | 1.257 | 1.272 |
| **Hispanic** |  |  |  |  |  |  |  |  |  |
| 2020 | 167,347 | 126,119 | 293,466 | 17,890,593 | 17,867,600 | 35,758,193 | 1,592 | 1,016 | 1,279 |
| 2019 | 109,426 | 90,347 | 199,773 | 17,491,893 | 17,462,061 | 34,953,954 | 1,106 | 762 | 918 |
| Absolute Change | 57,921 | 35,772 | 93,693 | 398,700 | 405,539 | 804,239 | 487 | 254 | 361 |
| Ratio | 1.529 | 1.396 | 1.469 | 1.023 | 1.023 | 1.023 | 1.440 | 1.334 | 1.393 |

**Table B.**
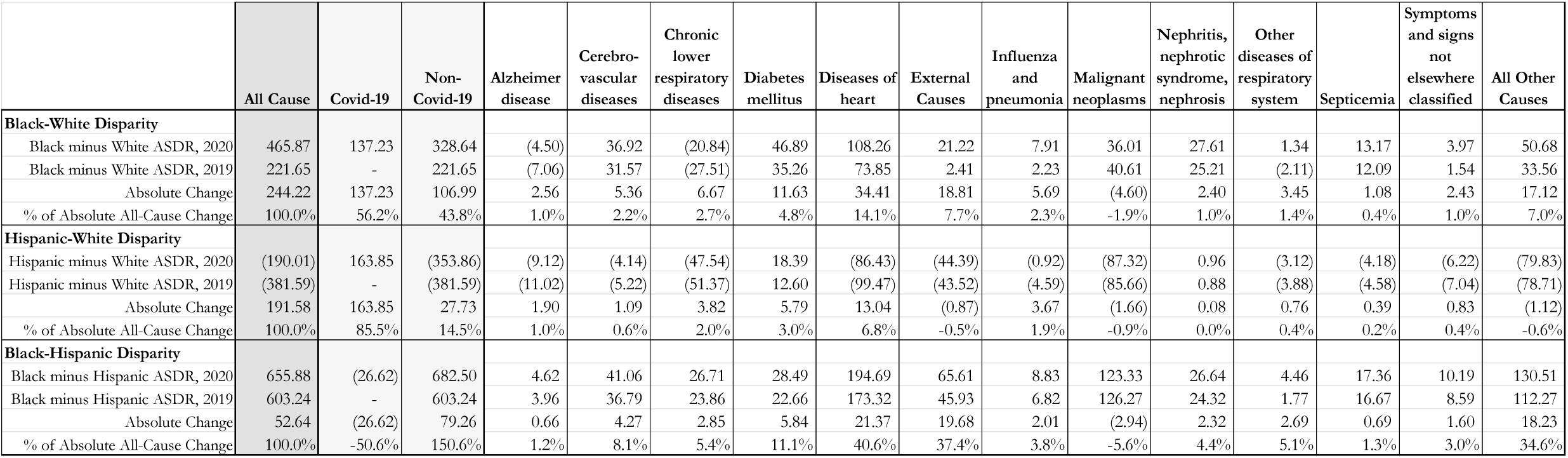
Change in ASDR (ages 25+) Racial Disparities by Cause of Death and Race/Ethnicity, Both Sexes.

**Table C.** Change in ASDR (ages 25-64 and ages 65+) by Cause of Death and Race/Ethnicity, Both Sexes.

|  | All Cause | Covid-19 | Non-Covid-19 | Alzheimer disease | Cerebro-vascular diseases | Chronic lower respiratory diseases | Diabetes mellitus | Diseases of heart | External Causes | Influenza and pneumonia | Malignant neoplasms | Nephritis, nephrotic syndrome, nephrosis | Other diseases of respiratory system | Septicemia | Symptoms and signs not elsewhere classified | All Other Causes |
| --- | --- | --- | --- | --- | --- | --- | --- | --- | --- | --- | --- | --- | --- | --- | --- | --- |
| **ASDR AGES 25 to 64** |  |  |  |  |  |  |  |  |  |  |  |  |  |  |  |  |
| **Non-Hispanic White** |  |  |  |  |  |  |  |  |  |  |  |  |  |  |  |  |
| ASDR, 2020 | 460.68 | 20.07 | 440.61 | 0.98 | 10.97 | 16.29 | 14.45 | 79.09 | 108.59 | 5.81 | 97.55 | 4.21 | 4.37 | 5.76 | 4.81 | 87.72 |
| ASDR, 2019 | 405.64 |  | 405.64 | 0.94 | 10.03 | 16.26 | 12.74 | 72.23 | 95.34 | 5.06 | 97.94 | 4.15 | 4.12 | 5.03 | 4.58 | 77.22 |
| % Change | 13.6% | 100.0% | 8.6% | 4.8% | 9.4% | 0.2% | 13.4% | 9.5% | 13.9% | 14.8% | -0.4% | 1.6% | 6.0% | 14.6% | 4.8% | 13.6% |
| Absolute Change | 55.03 | 20.07 | 34.97 | 0.05 | 0.94 | 0.03 | 1.70 | 6.87 | 13.24 | 0.75 | (0.38) | 0.07 | 0.25 | 0.73 | 0.22 | 10.50 |
| % of Absolute All-Cause Change | 100.0% | 36.5% | 63.5% | 0.1% | 1.7% | 0.1% | 3.1% | 12.5% | 24.1% | 1.4% | -0.7% | 0.1% | 0.5% | 1.3% | 0.4% | 19.1% |
| **Non-Hispanic Black** |  |  |  |  |  |  |  |  |  |  |  |  |  |  |  |  |
| ASDR, 2020 | 761.44 | 77.25 | 684.19 | 0.85 | 27.93 | 16.03 | 35.98 | 154.14 | 147.38 | 10.39 | 121.25 | 15.33 | 8.20 | 11.05 | 8.41 | 127.25 |
| ASDR, 2019 | 596.65 |  | 596.65 | 0.71 | 24.89 | 14.95 | 27.71 | 134.22 | 112.12 | 7.94 | 124.65 | 13.82 | 6.63 | 10.25 | 7.31 | 111.44 |
| % Change | 27.6% | 100.0% | 14.7% | 19.1% | 12.2% | 7.2% | 29.9% | 14.8% | 31.5% | 30.8% | -2.7% | 11.0% | 23.6% | 7.7% | 15.0% | 14.2% |
| Absolute Change | 164.79 | 77.25 | 87.54 | 0.14 | 3.05 | 1.08 | 8.27 | 19.91 | 35.26 | 2.45 | (3.40) | 1.51 | 1.57 | 0.79 | 1.09 | 15.81 |
| % of Absolute All-Cause Change | 100.0% | 46.9% | 53.1% | 0.1% | 1.8% | 0.7% | 5.0% | 12.1% | 21.4% | 1.5% | -2.1% | 0.9% | 1.0% | 0.5% | 0.7% | 9.6% |
| **Hispanic** |  |  |  |  |  |  |  |  |  |  |  |  |  |  |  |  |
| ASDR, 2020 | 405.83 | 94.72 | 311.11 | 0.66 | 11.81 | 4.02 | 17.50 | 51.32 | 70.31 | 5.56 | 64.73 | 5.44 | 3.82 | 4.14 | 3.47 | 68.32 |
| ASDR, 2019 | 275.86 |  | 275.86 | 0.59 | 10.55 | 3.64 | 14.22 | 44.87 | 58.69 | 4.06 | 65.92 | 4.98 | 3.08 | 3.71 | 2.96 | 58.59 |
| % Change | 47.1% | 100.0% | 12.8% | 11.9% | 12.0% | 10.3% | 23.1% | 14.4% | 19.8% | 36.9% | -1.8% | 9.3% | 23.9% | 11.9% | 17.2% | 16.6% |
| Absolute Change | 129.97 | 94.72 | 35.25 | 0.07 | 1.26 | 0.37 | 3.28 | 6.44 | 11.62 | 1.50 | (1.19) | 0.46 | 0.74 | 0.44 | 0.51 | 9.73 |
| % of Absolute All-Cause Change | 100.0% | 72.9% | 27.1% | 0.1% | 1.0% | 0.3% | 2.5% | 5.0% | 8.9% | 1.2% | -0.9% | 0.4% | 0.6% | 0.3% | 0.4% | 7.5% |
| **ASDR AGES 65+** |  |  |  |  |  |  |  |  |  |  |  |  |  |  |  |  |
| **Non-Hispanic White** |  |  |  |  |  |  |  |  |  |  |  |  |  |  |  |  |
| ASDR, 2020 | 4,566.02 | 434.89 | 4,131.13 | 249.95 | 242.22 | 262.19 | 112.08 | 1,027.54 | 151.32 | 75.37 | 826.34 | 70.50 | 69.50 | 52.49 | 42.39 | 949.23 |
| ASDR, 2019 | 4,046.41 |  | 4,046.41 | 231.48 | 233.61 | 279.31 | 102.06 | 1,015.26 | 150.36 | 75.81 | 834.99 | 71.80 | 71.23 | 53.13 | 42.51 | 884.87 |
| % Change | 12.8% | 100.0% | 2.1% | 8.0% | 3.7% | -6.1% | 9.8% | 1.2% | 0.6% | -0.6% | -1.0% | -1.8% | -2.4% | -1.2% | -0.3% | 7.3% |
| Absolute Change | 519.61 | 434.89 | 84.72 | 18.47 | 8.61 | (17.12) | 10.02 | 12.28 | 0.96 | (0.44) | (8.65) | (1.29) | (1.73) | (0.64) | (0.12) | 64.36 |
| % of Absolute All-Cause Change | 100.0% | 83.7% | 16.3% | 3.6% | 1.7% | -3.3% | 1.9% | 2.4% | 0.2% | -0.1% | -1.7% | -0.2% | -0.3% | -0.1% | 0.0% | 12.4% |
| **Non-Hispanic Black** |  |  |  |  |  |  |  |  |  |  |  |  |  |  |  |  |
| ASDR, 2020 | 5,539.08 | 818.00 | 4,721.08 | 232.04 | 340.47 | 178.12 | 236.86 | 1,237.84 | 118.55 | 93.51 | 900.19 | 148.74 | 63.20 | 89.90 | 47.51 | 1,034.16 |
| ASDR, 2019 | 4,362.19 |  | 4,362.19 | 203.42 | 316.52 | 171.32 | 199.67 | 1,125.54 | 108.65 | 76.02 | 918.28 | 144.72 | 54.93 | 86.32 | 40.41 | 916.40 |
| % Change | 27.0% | 100.0% | 8.2% | 14.1% | 7.6% | 4.0% | 18.6% | 10.0% | 9.1% | 23.0% | -2.0% | 2.8% | 15.0% | 4.1% | 17.5% | 12.9% |
| Absolute Change | 1,176.88 | 818.00 | 358.88 | 28.62 | 23.95 | 6.80 | 37.19 | 112.30 | 9.91 | 17.49 | (18.09) | 4.02 | 8.26 | 3.57 | 7.09 | 117.76 |
| % of Absolute All-Cause Change | 100.0% | 69.5% | 30.5% | 2.4% | 2.0% | 0.6% | 3.2% | 9.5% | 0.8% | 1.5% | -1.5% | 0.3% | 0.7% | 0.3% | 0.6% | 10.0% |
| **Hispanic** |  |  |  |  |  |  |  |  |  |  |  |  |  |  |  |  |
| ASDR, 2020 | 3,960.82 | 872.73 | 3,088.09 | 213.81 | 222.80 | 106.30 | 177.59 | 760.94 | 88.15 | 72.40 | 571.63 | 70.64 | 58.47 | 40.42 | 21.17 | 683.78 |
| ASDR, 2019 | 2,891.31 |  | 2,891.31 | 187.66 | 210.76 | 108.92 | 148.83 | 694.27 | 85.75 | 60.19 | 584.55 | 72.82 | 58.60 | 38.56 | 18.81 | 621.59 |
| % Change | 37.0% | 100.0% | 6.8% | 13.9% | 5.7% | -2.4% | 19.3% | 9.6% | 2.8% | 20.3% | -2.2% | -3.0% | -0.2% | 4.8% | 12.5% | 10.0% |
| Absolute Change | 1,069.51 | 872.73 | 196.78 | 26.15 | 12.04 | (2.61) | 28.75 | 66.67 | 2.40 | 12.21 | (12.92) | (2.19) | (0.13) | 1.86 | 2.36 | 62.19 |
| % of Absolute All-Cause Change | 100.0% | 81.6% | 18.4% | 2.4% | 1.1% | -0.2% | 2.7% | 6.2% | 0.2% | 1.1% | -1.2% | -0.2% | 0.0% | 0.2% | 0.2% | 5.8% |

**Table D.** Change in ASDR by Cause of Death and Race/Ethnicity, Male Population.

|  | All Cause | Covid-19 | Non-Covid-19 | Alzheimer disease | Cerebro-vascular diseases | Chronic lower respiratory diseases | Diabetes mellitus | Diseases of heart | External Causes | Influenza and pneumonia | Malignant neoplasms | Nephritis, nephrotic syndrome, nephrosis | Other diseases of respiratory system | Septicemia | Symptoms and signs not elsewhere classified | All Other Causes |
| --- | --- | --- | --- | --- | --- | --- | --- | --- | --- | --- | --- | --- | --- | --- | --- | --- |
| **ASDR AGES 25+** |  |  |  |  |  |  |  |  |  |  |  |  |  |  |  |  |
| **Non-Hispanic White** |  |  |  |  |  |  |  |  |  |  |  |  |  |  |  |  |
| ASDR, 2020 | 1,727.47 | 150.26 | 1,577.21 | 48.09 | 68.04 | 82.61 | 49.58 | 397.37 | 166.92 | 27.24 | 327.62 | 25.22 | 24.37 | 19.09 | 14.79 | 326.27 |
| ASDR, 2019 | 1,521.41 |  | 1,521.41 | 45.71 | 64.89 | 85.82 | 45.24 | 384.83 | 152.10 | 26.01 | 330.56 | 25.49 | 24.66 | 18.88 | 14.69 | 302.54 |
| % Change | 13.5% | 100.0% | 3.7% | 5.2% | 4.9% | -3.7% | 9.6% | 3.3% | 9.7% | 4.7% | -0.9% | -1.1% | -1.2% | 1.1% | 0.6% | 7.8% |
| Absolute Change | 206.05 | 150.26 | 55.79 | 2.38 | 3.16 | (3.21) | 4.34 | 12.54 | 14.82 | 1.23 | (2.94) | (0.27) | (0.29) | 0.21 | 0.09 | 23.73 |
| % of Absolute All-Cause Change | 100.0% | 72.9% | 27.1% | 1.2% | 1.5% | -1.6% | 2.1% | 6.1% | 7.2% | 0.6% | -1.4% | -0.1% | -0.1% | 0.1% | 0.0% | 11.5% |
| **Non-Hispanic Black** |  |  |  |  |  |  |  |  |  |  |  |  |  |  |  |  |
| ASDR, 2020 | 2,410.99 | 339.07 | 2,071.92 | 46.30 | 117.35 | 69.94 | 104.51 | 540.70 | 225.78 | 39.73 | 387.32 | 57.67 | 25.48 | 34.76 | 21.61 | 400.79 |
| ASDR, 2019 | 1,877.17 |  | 1,877.17 | 41.42 | 105.71 | 67.92 | 85.05 | 485.47 | 178.02 | 30.18 | 395.31 | 55.85 | 21.49 | 33.92 | 18.58 | 358.25 |
| % Change | 28.4% | 100.0% | 10.4% | 11.8% | 11.0% | 3.0% | 22.9% | 11.4% | 26.8% | 31.6% | -2.0% | 3.2% | 18.5% | 2.5% | 16.3% | 11.9% |
| Absolute Change | 533.82 | 339.07 | 194.75 | 4.88 | 11.64 | 2.02 | 19.46 | 55.24 | 47.75 | 9.55 | (7.99) | 1.81 | 3.98 | 0.84 | 3.02 | 42.54 |
| % of Absolute All-Cause Change | 100.0% | 63.5% | 36.5% | 0.9% | 2.2% | 0.4% | 3.6% | 10.3% | 8.9% | 1.8% | -1.5% | 0.3% | 0.7% | 0.2% | 0.6% | 8.0% |
| **Hispanic** |  |  |  |  |  |  |  |  |  |  |  |  |  |  |  |  |
| ASDR, 2020 | 1,592.46 | 394.17 | 1,198.29 | 42.15 | 68.82 | 35.73 | 69.43 | 289.71 | 114.78 | 27.34 | 224.51 | 26.02 | 20.78 | 14.77 | 9.61 | 254.65 |
| ASDR, 2019 | 1,105.90 |  | 1,105.90 | 37.63 | 64.56 | 36.47 | 57.76 | 260.98 | 98.78 | 21.73 | 231.72 | 25.90 | 19.19 | 14.57 | 8.52 | 228.11 |
| % Change | 44.0% | 100.0% | 8.4% | 12.0% | 6.6% | -2.0% | 20.2% | 11.0% | 16.2% | 25.8% | -3.1% | 0.4% | 8.3% | 1.4% | 12.7% | 11.6% |
| Absolute Change | 486.56 | 394.17 | 92.39 | 4.52 | 4.26 | (0.73) | 11.68 | 28.73 | 16.00 | 5.61 | (7.21) | 0.12 | 1.59 | 0.21 | 1.08 | 26.54 |
| % of Absolute All-Cause Change | 100.0% | 81.0% | 19.0% | 0.9% | 0.9% | -0.2% | 2.4% | 5.9% | 3.3% | 1.2% | -1.5% | 0.0% | 0.3% | 0.0% | 0.2% | 5.5% |
| **ASDR AGES 25 to 64** |  |  |  |  |  |  |  |  |  |  |  |  |  |  |  |  |
| **Non-Hispanic White** |  |  |  |  |  |  |  |  |  |  |  |  |  |  |  |  |
| ASDR, 2020 | 584.58 | 25.59 | 558.99 | 0.82 | 13.00 | 16.56 | 18.91 | 113.39 | 155.75 | 6.59 | 104.49 | 5.11 | 4.90 | 6.12 | 6.05 | 107.31 |
| ASDR, 2019 | 510.21 |  | 510.21 | 0.81 | 11.59 | 16.11 | 16.87 | 103.10 | 136.56 | 5.69 | 104.90 | 5.00 | 4.58 | 5.46 | 5.68 | 93.88 |
| % Change | 14.6% | 100.0% | 9.6% | 0.5% | 12.1% | 2.8% | 12.1% | 10.0% | 14.1% | 15.8% | -0.4% | 2.3% | 7.1% | 12.1% | 6.6% | 14.3% |
| Absolute Change | 74.37 | 25.59 | 48.78 | 0.00 | 1.41 | 0.45 | 2.04 | 10.29 | 19.19 | 0.90 | (0.41) | 0.11 | 0.33 | 0.66 | 0.37 | 13.44 |
| % of Absolute All-Cause Change | 100.0% | 34.4% | 65.6% | 0.0% | 1.9% | 0.6% | 2.7% | 13.8% | 25.8% | 1.2% | -0.5% | 0.2% | 0.4% | 0.9% | 0.5% | 18.1% |
| **Non-Hispanic Black** |  |  |  |  |  |  |  |  |  |  |  |  |  |  |  |  |
| ASDR, 2020 | 997.04 | 97.98 | 899.06 | 0.89 | 34.69 | 17.25 | 45.82 | 214.51 | 238.78 | 12.82 | 128.45 | 18.73 | 9.12 | 11.94 | 11.18 | 154.87 |
| ASDR, 2019 | 771.33 |  | 771.33 | 0.71 | 30.32 | 16.26 | 34.34 | 185.51 | 181.70 | 9.61 | 132.93 | 17.01 | 6.99 | 11.02 | 9.86 | 135.06 |
| % Change | 29.3% | 100.0% | 16.6% | 25.2% | 14.4% | 6.1% | 33.4% | 15.6% | 31.4% | 33.5% | -3.4% | 10.1% | 30.4% | 8.3% | 13.3% | 14.7% |
| Absolute Change | 225.70 | 97.98 | 127.73 | 0.18 | 4.37 | 1.00 | 11.48 | 29.00 | 57.08 | 3.21 | (4.48) | 1.72 | 2.13 | 0.91 | 1.31 | 19.81 |
| % of Absolute All-Cause Change | 100.0% | 43.4% | 56.6% | 0.1% | 1.9% | 0.4% | 5.1% | 12.8% | 25.3% | 1.4% | -2.0% | 0.8% | 0.9% | 0.4% | 0.6% | 8.8% |
| **Hispanic** |  |  |  |  |  |  |  |  |  |  |  |  |  |  |  |  |
| ASDR, 2020 | 552.41 | 135.39 | 417.02 | 0.59 | 15.27 | 4.31 | 22.77 | 76.31 | 111.08 | 7.22 | 66.19 | 6.90 | 4.62 | 4.86 | 5.11 | 91.78 |
| ASDR, 2019 | 363.78 |  | 363.78 | 0.53 | 13.45 | 4.07 | 18.33 | 66.66 | 91.73 | 5.07 | 67.42 | 6.28 | 3.51 | 4.62 | 4.36 | 77.74 |
| % Change | 51.9% | 100.0% | 14.6% | 11.4% | 13.5% | 6.0% | 24.2% | 14.5% | 21.1% | 42.3% | -1.8% | 10.0% | 31.7% | 5.1% | 17.3% | 18.1% |
| Absolute Change | 188.64 | 135.39 | 53.24 | 0.06 | 1.81 | 0.24 | 4.44 | 9.65 | 19.36 | 2.15 | (1.23) | 0.63 | 1.11 | 0.24 | 0.76 | 14.04 |
| % of Absolute All-Cause Change | 100.0% | 71.8% | 28.2% | 0.0% | 1.0% | 0.1% | 2.4% | 5.1% | 10.3% | 1.1% | -0.7% | 0.3% | 0.6% | 0.1% | 0.4% | 7.4% |
| **ASDR AGES 65+** |  |  |  |  |  |  |  |  |  |  |  |  |  |  |  |  |
| **Non-Hispanic White** |  |  |  |  |  |  |  |  |  |  |  |  |  |  |  |  |
| ASDR, 2020 | 5,238.22 | 533.24 | 4,704.98 | 193.30 | 237.15 | 285.52 | 143.80 | 1,269.72 | 201.24 | 90.68 | 1,013.01 | 86.99 | 84.16 | 58.91 | 41.63 | 998.88 |
| ASDR, 2019 | 4,627.66 |  | 4,627.66 | 183.61 | 228.60 | 299.98 | 132.40 | 1,250.27 | 199.85 | 88.43 | 1,023.73 | 88.45 | 86.35 | 60.08 | 42.40 | 943.52 |
| % Change | 13.2% | 100.0% | 1.7% | 5.3% | 3.7% | -4.8% | 8.6% | 1.6% | 0.7% | 2.5% | -1.0% | -1.7% | -2.5% | -2.0% | -1.8% | 5.9% |
| Absolute Change | 610.56 | 533.24 | 77.32 | 9.70 | 8.54 | (14.46) | 11.41 | 19.45 | 1.39 | 2.25 | (10.72) | (1.46) | (2.19) | (1.17) | (0.77) | 55.36 |
| % of Absolute All-Cause Change | 100.0% | 87.3% | 12.7% | 1.6% | 1.4% | -2.4% | 1.9% | 3.2% | 0.2% | 0.4% | -1.8% | -0.2% | -0.4% | -0.2% | -0.1% | 9.1% |
| **Non-Hispanic Black** |  |  |  |  |  |  |  |  |  |  |  |  |  |  |  |  |
| ASDR, 2020 | 6,754.42 | 1,079.67 | 5,674.76 | 185.76 | 371.28 | 231.80 | 284.79 | 1,542.71 | 185.83 | 122.38 | 1,182.51 | 177.26 | 75.71 | 104.87 | 53.65 | 1,156.22 |
| ASDR, 2019 | 5,274.13 |  | 5,274.13 | 166.44 | 337.29 | 226.63 | 240.81 | 1,406.88 | 166.73 | 93.38 | 1,201.29 | 175.16 | 66.03 | 104.24 | 45.38 | 1,043.87 |
| % Change | 28.1% | 100.0% | 7.6% | 11.6% | 10.1% | 2.3% | 18.3% | 9.7% | 11.5% | 31.1% | -1.6% | 1.2% | 14.7% | 0.6% | 18.2% | 10.8% |
| Absolute Change | 1,480.30 | 1,079.67 | 400.63 | 19.32 | 33.99 | 5.18 | 43.98 | 135.83 | 19.10 | 29.00 | (18.79) | 2.10 | 9.68 | 0.63 | 8.27 | 112.35 |
| % of Absolute All-Cause Change | 100.0% | 72.9% | 27.1% | 1.3% | 2.3% | 0.3% | 3.0% | 9.2% | 1.3% | 2.0% | -1.3% | 0.1% | 0.7% | 0.0% | 0.6% | 7.6% |
| **Hispanic** |  |  |  |  |  |  |  |  |  |  |  |  |  |  |  |  |
| ASDR, 2020 | 4,787.31 | 1,189.08 | 3,598.23 | 169.80 | 233.31 | 132.24 | 212.76 | 945.23 | 126.15 | 89.17 | 710.83 | 84.73 | 70.41 | 45.23 | 23.41 | 754.95 |
| ASDR, 2019 | 3,385.57 |  | 3,385.57 | 151.59 | 221.55 | 135.98 | 178.86 | 857.86 | 120.44 | 72.92 | 736.42 | 86.18 | 67.35 | 45.11 | 21.32 | 689.99 |
| % Change | 41.4% | 100.0% | 6.3% | 12.0% | 5.3% | -2.7% | 19.0% | 10.2% | 4.7% | 22.3% | -3.5% | -1.7% | 4.5% | 0.3% | 9.8% | 9.4% |
| Absolute Change | 1,401.73 | 1,189.08 | 212.66 | 18.21 | 11.76 | (3.74) | 33.91 | 87.37 | 5.71 | 16.25 | (25.59) | (1.45) | 3.06 | 0.12 | 2.09 | 64.95 |
| % of Absolute All-Cause Change | 100.0% | 84.8% | 15.2% | 1.3% | 0.8% | -0.3% | 2.4% | 6.2% | 0.4% | 1.2% | -1.8% | -0.1% | 0.2% | 0.0% | 0.1% | 4.6% |

**Table E.** Change in ASDR by Cause of Death and Race/Ethnicity, Female Population.

|  | All Cause | Covid-19 | Non-Covid-19 | Alzheimer disease | Cerebro-vascular diseases | Chronic lower respiratory diseases | Diabetes mellitus | Diseases of heart | External Causes | Influenza and pneumonia | Malignant neoplasms | Nephritis, nephrotic syndrome, nephrosis | Other diseases of respiratory system | Septicemia | Symptoms and signs not elsewhere classified | All Other Causes |
| --- | --- | --- | --- | --- | --- | --- | --- | --- | --- | --- | --- | --- | --- | --- | --- | --- |
| **ASDR AGES 25+** |  |  |  |  |  |  |  |  |  |  |  |  |  |  |  |  |
| **Non-Hispanic White** |  |  |  |  |  |  |  |  |  |  |  |  |  |  |  |  |
| ASDR, 2020 | 1,245.15 | 99.35 | 1,145.80 | 71.21 | 66.42 | 72.16 | 29.05 | 240.50 | 73.69 | 19.62 | 236.99 | 16.98 | 17.24 | 15.81 | 13.04 | 273.11 |
| ASDR, 2019 | 1,109.02 |  | 1,109.02 | 65.10 | 63.90 | 77.24 | 25.84 | 236.49 | 68.13 | 19.72 | 239.10 | 17.27 | 17.50 | 15.30 | 12.94 | 250.49 |
| % Change | 12.3% | 100.0% | 3.3% | 9.4% | 3.9% | -6.6% | 12.4% | 1.7% | 8.2% | -0.5% | -0.9% | -1.7% | -1.5% | 3.3% | 0.8% | 9.0% |
| Absolute Change | 136.13 | 99.35 | 36.78 | 6.10 | 2.52 | (5.08) | 3.21 | 4.01 | 5.56 | (0.09) | (2.11) | (0.30) | (0.27) | 0.51 | 0.10 | 22.62 |
| % of Absolute All-Cause Change | 100.0% | 73.0% | 27.0% | 4.5% | 1.9% | -3.7% | 2.4% | 2.9% | 4.1% | -0.1% | -1.6% | -0.2% | -0.2% | 0.4% | 0.1% | 16.6% |
| **Non-Hispanic Black** |  |  |  |  |  |  |  |  |  |  |  |  |  |  |  |  |
| ASDR, 2020 | 1,580.79 | 205.12 | 1,375.67 | 63.09 | 94.21 | 46.82 | 71.17 | 330.87 | 66.11 | 24.84 | 265.13 | 41.51 | 19.12 | 27.53 | 14.99 | 310.29 |
| ASDR, 2019 | 1,257.78 |  | 1,257.78 | 54.86 | 88.47 | 44.34 | 58.84 | 298.47 | 53.93 | 21.01 | 271.18 | 39.30 | 16.62 | 25.68 | 12.80 | 272.28 |
| % Change | 25.7% | 100.0% | 9.4% | 15.0% | 6.5% | 5.6% | 21.0% | 10.9% | 22.6% | 18.2% | -2.2% | 5.6% | 15.0% | 7.2% | 17.1% | 14.0% |
| Absolute Change | 323.01 | 205.12 | 117.89 | 8.23 | 5.73 | 2.48 | 12.33 | 32.40 | 12.18 | 3.83 | (6.05) | 2.21 | 2.50 | 1.85 | 2.19 | 38.01 |
| % of Absolute All-Cause Change | 100.0% | 63.5% | 36.5% | 2.5% | 1.8% | 0.8% | 3.8% | 10.0% | 3.8% | 1.2% | -1.9% | 0.7% | 0.8% | 0.6% | 0.7% | 11.8% |
| **Hispanic** |  |  |  |  |  |  |  |  |  |  |  |  |  |  |  |  |
| ASDR, 2020 | 1,016.13 | 198.05 | 818.07 | 59.66 | 58.63 | 24.54 | 46.33 | 173.27 | 35.75 | 17.88 | 163.70 | 17.94 | 14.55 | 11.68 | 6.11 | 188.02 |
| ASDR, 2019 | 761.86 |  | 761.86 | 51.91 | 55.22 | 24.65 | 38.67 | 158.69 | 33.29 | 14.83 | 165.75 | 18.33 | 14.87 | 10.49 | 5.29 | 169.87 |
| % Change | 33.4% | 100.0% | 7.4% | 14.9% | 6.2% | -0.4% | 19.8% | 9.2% | 7.4% | 20.6% | -1.2% | -2.1% | -2.1% | 11.4% | 15.3% | 10.7% |
| Absolute Change | 254.27 | 198.05 | 56.22 | 7.76 | 3.41 | (0.11) | 7.66 | 14.58 | 2.46 | 3.06 | (2.05) | (0.39) | (0.32) | 1.20 | 0.81 | 18.16 |
| % of Absolute All-Cause Change | 100.0% | 77.9% | 22.1% | 3.1% | 1.3% | 0.0% | 3.0% | 5.7% | 1.0% | 1.2% | -0.8% | -0.2% | -0.1% | 0.5% | 0.3% | 7.1% |
| **ASDR AGES 25 to 64** |  |  |  |  |  |  |  |  |  |  |  |  |  |  |  |  |
| **Non-Hispanic White** |  |  |  |  |  |  |  |  |  |  |  |  |  |  |  |  |
| ASDR, 2020 | 338.87 | 14.72 | 324.14 | 1.14 | 9.01 | 16.04 | 10.10 | 45.77 | 61.24 | 5.06 | 90.98 | 3.34 | 3.85 | 5.41 | 3.58 | 68.61 |
| ASDR, 2019 | 302.98 |  | 302.98 | 1.05 | 8.53 | 16.42 | 8.73 | 42.25 | 54.05 | 4.46 | 91.35 | 3.32 | 3.68 | 4.61 | 3.51 | 61.00 |
| % Change | 11.8% | 100.0% | 7.0% | 8.1% | 5.6% | -2.3% | 15.7% | 8.3% | 13.3% | 13.4% | -0.4% | 0.5% | 4.8% | 17.4% | 2.0% | 12.5% |
| Absolute Change | 35.89 | 14.72 | 21.16 | 0.09 | 0.48 | (0.38) | 1.37 | 3.52 | 7.19 | 0.60 | (0.37) | 0.02 | 0.17 | 0.80 | 0.07 | 7.61 |
| % of Absolute All-Cause Change | 100.0% | 41.0% | 59.0% | 0.2% | 1.3% | -1.1% | 3.8% | 9.8% | 20.0% | 1.7% | -1.0% | 0.0% | 0.5% | 2.2% | 0.2% | 21.2% |
| **Non-Hispanic Black** |  |  |  |  |  |  |  |  |  |  |  |  |  |  |  |  |
| ASDR, 2020 | 553.82 | 59.38 | 494.44 | 0.80 | 22.14 | 14.98 | 27.46 | 102.02 | 63.87 | 8.31 | 115.36 | 12.40 | 7.41 | 10.30 | 5.97 | 103.41 |
| ASDR, 2019 | 443.05 |  | 443.05 | 0.71 | 20.23 | 13.84 | 21.94 | 89.95 | 48.86 | 6.51 | 117.84 | 11.07 | 6.32 | 9.60 | 5.07 | 91.11 |
| % Change | 25.0% | 100.0% | 11.6% | 13.9% | 9.4% | 8.2% | 25.1% | 13.4% | 30.7% | 27.7% | -2.1% | 12.1% | 17.2% | 7.3% | 17.8% | 13.5% |
| Absolute Change | 110.77 | 59.38 | 51.39 | 0.10 | 1.91 | 1.14 | 5.52 | 12.07 | 15.01 | 1.80 | (2.48) | 1.33 | 1.09 | 0.70 | 0.90 | 12.30 |
| % of Absolute All-Cause Change | 100.0% | 53.6% | 46.4% | 0.1% | 1.7% | 1.0% | 5.0% | 10.9% | 13.6% | 1.6% | -2.2% | 1.2% | 1.0% | 0.6% | 0.8% | 11.1% |
| **Hispanic** |  |  |  |  |  |  |  |  |  |  |  |  |  |  |  |  |
| ASDR, 2020 | 259.20 | 54.59 | 204.61 | 0.73 | 8.39 | 3.73 | 12.32 | 26.70 | 28.06 | 3.92 | 63.49 | 4.02 | 3.03 | 3.43 | 1.80 | 44.98 |
| ASDR, 2019 | 188.01 |  | 188.01 | 0.65 | 7.71 | 3.22 | 10.19 | 23.51 | 24.59 | 3.06 | 64.67 | 3.73 | 2.67 | 2.81 | 1.55 | 39.65 |
| % Change | 37.9% | 100.0% | 8.8% | 12.8% | 8.9% | 15.6% | 20.9% | 13.6% | 14.1% | 28.2% | -1.8% | 7.8% | 13.7% | 22.1% | 16.2% | 13.4% |
| Absolute Change | 71.19 | 54.59 | 16.60 | 0.08 | 0.69 | 0.50 | 2.13 | 3.19 | 3.47 | 0.86 | (1.19) | 0.29 | 0.36 | 0.62 | 0.25 | 5.33 |
| % of Absolute All-Cause Change | 100.0% | 76.7% | 23.3% | 0.1% | 1.0% | 0.7% | 3.0% | 4.5% | 4.9% | 1.2% | -1.7% | 0.4% | 0.5% | 0.9% | 0.4% | 7.5% |
| **ASDR AGES 65+** |  |  |  |  |  |  |  |  |  |  |  |  |  |  |  |  |
| **Non-Hispanic White** |  |  |  |  |  |  |  |  |  |  |  |  |  |  |  |  |
| ASDR, 2020 | 4,029.10 | 359.29 | 3,669.81 | 286.44 | 242.76 | 244.54 | 87.24 | 838.66 | 111.93 | 64.38 | 685.49 | 58.87 | 58.35 | 47.74 | 42.13 | 901.28 |
| ASDR, 2019 | 3,585.05 |  | 3,585.05 | 261.85 | 233.97 | 264.08 | 78.37 | 833.17 | 111.38 | 66.59 | 692.94 | 60.13 | 59.97 | 48.14 | 41.91 | 832.55 |
| % Change | 12.4% | 100.0% | 2.4% | 9.4% | 3.8% | -7.4% | 11.3% | 0.7% | 0.5% | -3.3% | -1.1% | -2.1% | -2.7% | -0.8% | 0.5% | 8.3% |
| Absolute Change | 444.05 | 359.29 | 84.76 | 24.59 | 8.79 | (19.53) | 8.87 | 5.49 | 0.55 | (2.21) | (7.45) | (1.26) | (1.62) | (0.40) | 0.21 | 68.73 |
| % of Absolute All-Cause Change | 100.0% | 80.9% | 19.1% | 5.5% | 2.0% | -4.4% | 2.0% | 1.2% | 0.1% | -0.5% | -1.7% | -0.3% | -0.4% | -0.1% | 0.0% | 15.5% |
| **Non-Hispanic Black** |  |  |  |  |  |  |  |  |  |  |  |  |  |  |  |  |
| ASDR, 2020 | 4,735.48 | 652.79 | 4,082.68 | 254.42 | 315.57 | 144.65 | 205.43 | 1,033.86 | 73.00 | 75.59 | 725.21 | 130.91 | 55.09 | 80.47 | 42.68 | 945.79 |
| ASDR, 2019 | 3,760.49 |  | 3,760.49 | 221.23 | 298.11 | 138.03 | 172.18 | 939.01 | 69.51 | 65.54 | 742.20 | 126.02 | 48.25 | 75.07 | 36.54 | 828.80 |
| % Change | 25.9% | 100.0% | 8.6% | 15.0% | 5.9% | 4.8% | 19.3% | 10.1% | 5.0% | 15.3% | -2.3% | 3.9% | 14.2% | 7.2% | 16.8% | 14.1% |
| Absolute Change | 974.99 | 652.79 | 322.19 | 33.19 | 17.47 | 6.62 | 33.25 | 94.86 | 3.49 | 10.05 | (16.99) | 4.90 | 6.83 | 5.40 | 6.14 | 116.99 |
| % of Absolute All-Cause Change | 100.0% | 67.0% | 33.0% | 3.4% | 1.8% | 0.7% | 3.4% | 9.7% | 0.4% | 1.0% | -1.7% | 0.5% | 0.7% | 0.6% | 0.6% | 12.0% |
| **Hispanic** |  |  |  |  |  |  |  |  |  |  |  |  |  |  |  |  |
| ASDR, 2020 | 3,341.27 | 638.75 | 2,702.52 | 240.70 | 212.95 | 88.46 | 150.79 | 623.52 | 59.38 | 60.77 | 471.53 | 60.71 | 49.93 | 37.03 | 19.33 | 627.42 |
| ASDR, 2019 | 2,524.61 |  | 2,524.61 | 209.37 | 201.19 | 90.45 | 126.15 | 573.94 | 60.03 | 50.97 | 476.24 | 63.19 | 52.35 | 34.07 | 16.79 | 569.87 |
| % Change | 32.3% | 100.0% | 7.0% | 15.0% | 5.8% | -2.2% | 19.5% | 8.6% | -1.1% | 19.2% | -1.0% | -3.9% | -4.6% | 8.7% | 15.1% | 10.1% |
| Absolute Change | 816.65 | 638.75 | 177.91 | 31.33 | 11.76 | (1.99) | 24.64 | 49.57 | (0.64) | 9.80 | (4.71) | (2.48) | (2.42) | 2.96 | 2.54 | 57.55 |
| % of Absolute All-Cause Change | 100.0% | 78.2% | 21.8% | 3.8% | 1.4% | -0.2% | 3.0% | 6.1% | -0.1% | 1.2% | -0.6% | -0.3% | -0.3% | 0.4% | 0.3% | 7.0% |

**Table F.**
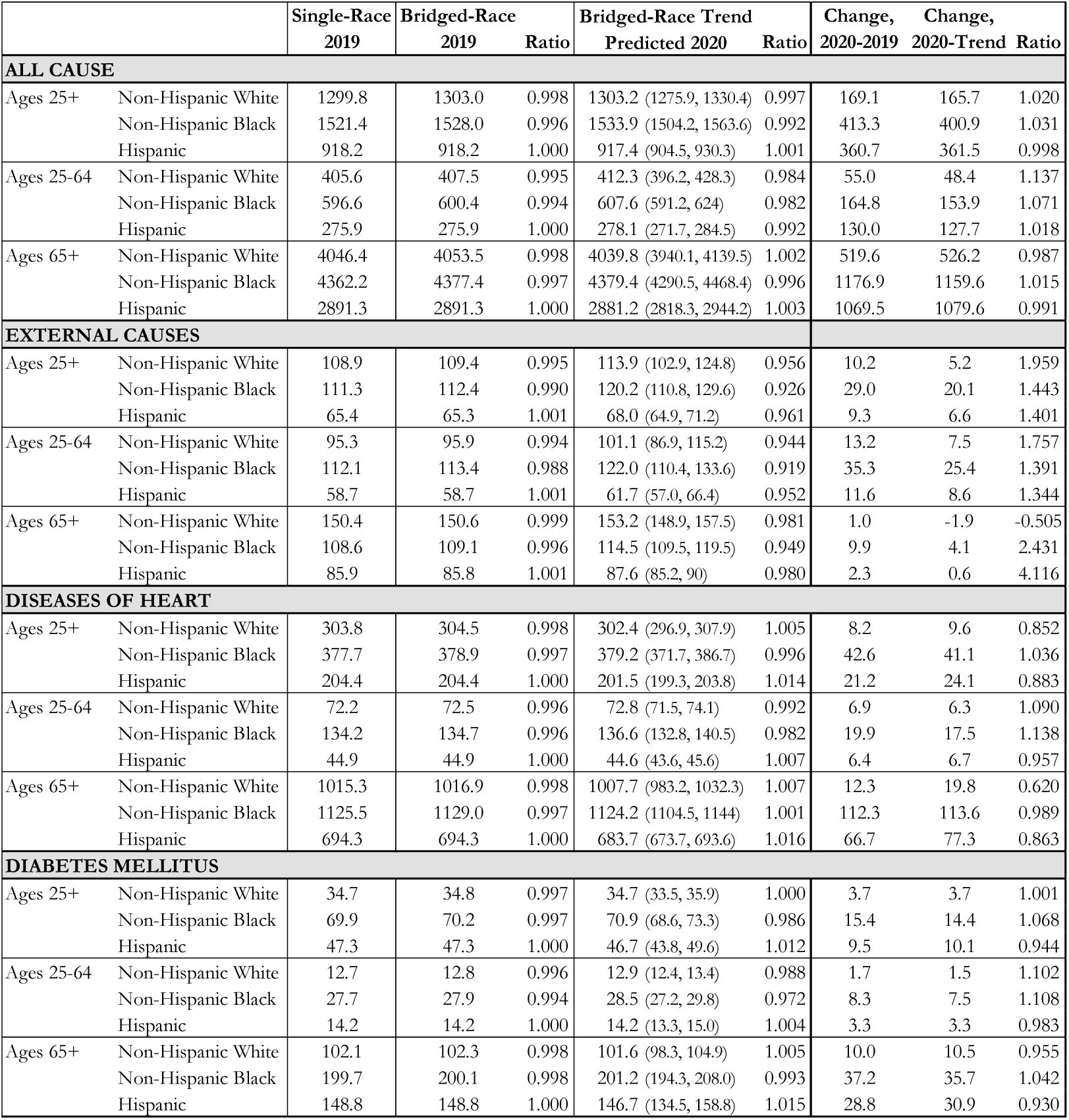
Sensitivity Analysis for Select Causes of Death.

|  |  | Single-Race<br>2019 | Bridged-Race<br>2019 | Ratio | Bridged-Race Trend<br>Predicted 2020 | Ratio | Change,<br>2020-2019 | Change,<br>2020-Trend | Ratio |
| --- | --- | --- | --- | --- | --- | --- | --- | --- | --- |
| **ALL CAUSE** |  |  |  |  |  |  |  |  |  |
| Ages 25+ | Non-Hispanic White | 1299.8 | 1303.0 | 0.998 | 1303.2 (1275.9, 1330.4) | 0.997 | 169.1 | 165.7 | 1.020 |
|  | Non-Hispanic Black | 1521.4 | 1528.0 | 0.996 | 1533.9 (1504.2, 1563.6) | 0.992 | 413.3 | 400.9 | 1.031 |
|  | Hispanic | 918.2 | 918.2 | 1.000 | 917.4 (904.5, 930.3) | 1.001 | 360.7 | 361.5 | 0.998 |
| Ages 25-64 | Non-Hispanic White | 405.6 | 407.5 | 0.995 | 412.3 (396.2, 428.3) | 0.984 | 55.0 | 48.4 | 1.137 |
|  | Non-Hispanic Black | 596.6 | 600.4 | 0.994 | 607.6 (591.2, 624) | 0.982 | 164.8 | 153.9 | 1.071 |
|  | Hispanic | 275.9 | 275.9 | 1.000 | 278.1 (271.7, 284.5) | 0.992 | 130.0 | 127.7 | 1.018 |
| Ages 65+ | Non-Hispanic White | 4046.4 | 4053.5 | 0.998 | 4039.8 (3940.1, 4139.5) | 1.002 | 519.6 | 526.2 | 0.987 |
|  | Non-Hispanic Black | 4362.2 | 4377.4 | 0.997 | 4379.4 (4290.5, 4468.4) | 0.996 | 1176.9 | 1159.6 | 1.015 |
|  | Hispanic | 2891.3 | 2891.3 | 1.000 | 2881.2 (2818.3, 2944.2) | 1.003 | 1069.5 | 1079.6 | 0.991 |
| **EXTERNAL CAUSES** |  |  |  |  |  |  |  |  |  |
| Ages 25+ | Non-Hispanic White | 108.9 | 109.4 | 0.995 | 113.9 (102.9, 124.8) | 0.956 | 10.2 | 5.2 | 1.959 |
|  | Non-Hispanic Black | 111.3 | 112.4 | 0.990 | 120.2 (110.8, 129.6) | 0.926 | 29.0 | 20.1 | 1.443 |
|  | Hispanic | 65.4 | 65.3 | 1.001 | 68.0 (64.9, 71.2) | 0.961 | 9.3 | 6.6 | 1.401 |
| Ages 25-64 | Non-Hispanic White | 95.3 | 95.9 | 0.994 | 101.1 (86.9, 115.2) | 0.944 | 13.2 | 7.5 | 1.757 |
|  | Non-Hispanic Black | 112.1 | 113.4 | 0.988 | 122.0 (110.4, 133.6) | 0.919 | 35.3 | 25.4 | 1.391 |
|  | Hispanic | 58.7 | 58.7 | 1.001 | 61.7 (57.0, 66.4) | 0.952 | 11.6 | 8.6 | 1.344 |
| Ages 65+ | Non-Hispanic White | 150.4 | 150.6 | 0.999 | 153.2 (148.9, 157.5) | 0.981 | 1.0 | -1.9 | -0.505 |
|  | Non-Hispanic Black | 108.6 | 109.1 | 0.996 | 114.5 (109.5, 119.5) | 0.949 | 9.9 | 4.1 | 2.431 |
|  | Hispanic | 85.9 | 85.8 | 1.001 | 87.6 (85.2, 90) | 0.980 | 2.3 | 0.6 | 4.116 |
| **DISEASES OF HEART** |  |  |  |  |  |  |  |  |  |
| Ages 25+ | Non-Hispanic White | 303.8 | 304.5 | 0.998 | 302.4 (296.9, 307.9) | 1.005 | 8.2 | 9.6 | 0.852 |
|  | Non-Hispanic Black | 377.7 | 378.9 | 0.997 | 379.2 (371.7, 386.7) | 0.996 | 42.6 | 41.1 | 1.036 |
|  | Hispanic | 204.4 | 204.4 | 1.000 | 201.5 (199.3, 203.8) | 1.014 | 21.2 | 24.1 | 0.883 |
| Ages 25-64 | Non-Hispanic White | 72.2 | 72.5 | 0.996 | 72.8 (71.5, 74.1) | 0.992 | 6.9 | 6.3 | 1.090 |
|  | Non-Hispanic Black | 134.2 | 134.7 | 0.996 | 136.6 (132.8, 140.5) | 0.982 | 19.9 | 17.5 | 1.138 |
|  | Hispanic | 44.9 | 44.9 | 1.000 | 44.6 (43.6, 45.6) | 1.007 | 6.4 | 6.7 | 0.957 |
| Ages 65+ | Non-Hispanic White | 1015.3 | 1016.9 | 0.998 | 1007.7 (983.2, 1032.3) | 1.007 | 12.3 | 19.8 | 0.620 |
|  | Non-Hispanic Black | 1125.5 | 1129.0 | 0.997 | 1124.2 (1104.5, 1144) | 1.001 | 112.3 | 113.6 | 0.989 |
|  | Hispanic | 694.3 | 694.3 | 1.000 | 683.7 (673.7, 693.6) | 1.016 | 66.7 | 77.3 | 0.863 |
| **DIABETES MELLITUS** |  |  |  |  |  |  |  |  |  |
| Ages 25+ | Non-Hispanic White | 34.7 | 34.8 | 0.997 | 34.7 (33.5, 35.9) | 1.000 | 3.7 | 3.7 | 1.001 |
|  | Non-Hispanic Black | 69.9 | 70.2 | 0.997 | 70.9 (68.6, 73.3) | 0.986 | 15.4 | 14.4 | 1.068 |
|  | Hispanic | 47.3 | 47.3 | 1.000 | 46.7 (43.8, 49.6) | 1.012 | 9.5 | 10.1 | 0.944 |
| Ages 25-64 | Non-Hispanic White | 12.7 | 12.8 | 0.996 | 12.9 (12.4, 13.4) | 0.988 | 1.7 | 1.5 | 1.102 |
|  | Non-Hispanic Black | 27.7 | 27.9 | 0.994 | 28.5 (27.2, 29.8) | 0.972 | 8.3 | 7.5 | 1.108 |
|  | Hispanic | 14.2 | 14.2 | 1.000 | 14.2 (13.3, 15.0) | 1.004 | 3.3 | 3.3 | 0.983 |
| Ages 65+ | Non-Hispanic White | 102.1 | 102.3 | 0.998 | 101.6 (98.3, 104.9) | 1.005 | 10.0 | 10.5 | 0.955 |
|  | Non-Hispanic Black | 199.7 | 200.1 | 0.998 | 201.2 (194.3, 208.0) | 0.993 | 37.2 | 35.7 | 1.042 |
|  | Hispanic | 148.8 | 148.8 | 1.000 | 146.7 (134.5, 158.8) | 1.015 | 28.8 | 30.9 | 0.930 |

## Background

Our primary analysis is based on a comparison of mortality in 2019 to that in 2020. An alternative approach would compare reported mortality in 2020 to predicted mortality in 2020, with the prediction based on death rates for some period prior to 2020.^12,18^ This approach is not feasible because the race-specific death data in 2020 is based on “single race” coding and such data are not available prior to 2018. However, we tested an alternative death series based on “bridged-race” deaths to investigate how mortality in 2019 would compare to projected mortality in 2020 based on a time series approach using data from 2015 to 2019. [Note that a bridged-race death series is not available in 2020.] The two left-most columns in Table F show that the ASDRs in 2019 using bridged-race deaths are within 1% of those based on single-race deaths.

## Methods

To develop our estimates of predicted mortality in 2020 based on bridged-race deaths, we use OLS regression applied to a time series of ASDRs from 2015 to 2019. We then extrapolate the series and produce an expected mortality level for each racial/ethnic group in 2020 [Table F]. This extrapolation was conducted for all-cause mortality and for the underlying causes of death that contributed most to overall mortality change (external causes, heart disease, and diabetes).

## Results

A comparison of the 2019 bridged-race ASDRs to those predicted by the time series approach shows that nearly all overall ASDRs in 2019 for each racial/ethnic and age-standardized broad age group are within a 95% confidence interval of the expected 2020 overall ASDR based on the prior 5-year trend. Additionally, at ages 25+ and 65+, all-cause ASDR values of 2019 mortality and projected 2020 mortality are within 1% of each other for all three racial/ethnic groups. At ages 25-64, however, predicted all-cause values for all three groups are above values in 2019 by 0.8%-1.6%. For heart disease and diabetes at ages 25+ and 65+, predicted and actual values are within 1.6% of each other for all three racial/ethnic groups. For Hispanic individuals in these categories, predicted values are 1.2%-1.5% lower than actual 2019 values.

The main exceptions to the similarity of actual and predicted values pertain to external causes and ages 25-64. Actual 2019 mortality is below predicted 2020 mortality for external causes in all racial/ethnic categories and all age intervals, reaching a maximum of 9.1% for Black individuals at ages 25-64. The discrepancy reflects a rising 5-year trend in mortality from external causes which, had it continued into 2020, would have produced higher mortality than observed in 2019. If we had used the projected value of external causes rather than the actual value in 2019, we would have projected less discrepancy between actual and expected mortality in 2020, shown in the three right-hand columns of Table F. While Black populations would continue to have the largest increase in external cause mortality in these age intervals, the increase at ages 25-64 would have been only 25.4/100,000 rather than 35.3. We conclude that our estimates of changes in mortality from external causes associated with the Covid-19 epidemic are sensitive to whether prior mortality trends are incorporated into estimates of non-Covid-19 mortality in 2020. To a lesser extent, such sensitivity also applies to mortality at ages 25-64 from all causes and from heart disease and diabetes. At ages 25+ and 65+ for causes of death other than external causes, results in this paper do not appear to be sensitive to whether or not mortality trends during 2015-19 are incorporated into expectations for mortality in 2020.

## Notes

### Competing Interest Statement

The authors have declared no competing interest.

### Funding Statement

The paper was supported by grants from the Robert Wood Johnson Foundation (#77521) and the National Institute on Aging (#R01-AG060115).

### Author Declarations

The analyses are based solely on de-identified publicly available secondary data.

## CITATIONS

1. Alsan M, Chandra A, Simon KI. The Great Unequalizer: Initial Health Effects of COVID-19 in the United States. National Bureau of Economic Research; 2021. doi:10.3386/w28958

2. Andrasfay T, Goldman N. Association of the COVID-19 Pandemic With Estimated Life Expectancy by Race/Ethnicity in the United States, 2020. JAMA Netw Open. 2021;4(6):e2114520. doi:10.1001/jamanetworkopen.2021.14520

3. Woolf SH, Masters RK, Aron LY. Effect of the covid-19 pandemic in 2020 on life expectancy across populations in the USA and other high income countries: simulations of provisional mortality data. BMJ. 2021;373:n1343. doi:10.1136/bmj.n1343

4. Lopez L, Hart LH, Katz MH. Racial and Ethnic Health Disparities Related to COVID-19. JAMA. 2021;325(8):719. doi:10.1001/jama.2020.26443

5. Macias Gil R, Marcelin JR, Zuniga-Blanco B, Marquez C, Mathew T, Piggott DA. COVID-19 Pandemic: Disparate Health Impact on the Hispanic/Latinx Population in the United States. J Infect Dis. 2020;222(10):1592–1595. doi:10.1093/infdis/jiaa474

6. Bailey ZD, Feldman JM, Bassett MT. How Structural Racism Works — Racist Policies as a Root Cause of U.S. Racial Health Inequities. N Engl J Med. 2021;384(8):768–773. doi:10.1056/NEJMms2025396

7. McClure ES, Vasudevan P, Bailey Z, Patel S, Robinson WR. Racial Capitalism Within Public Health-How Occupational Settings Drive COVID-19 Disparities. Am J Epidemiol. 2020;189(11):1244–1253. doi:10.1093/aje/kwaa126

8. Stokes AC, Lundberg DJ, Bor J, Elo IT, Hempstead K, Preston SH. Association of Health Care Factors with Excess Deaths Not Assigned to COVID-19 in the US. JAMA Netw Open. Published online 2021. doi:Forthcoming

9. Cantor JH, Sood N, Bravata D, Pera M, Whaley CM. The Impact of the COVID-19 Pandemic and Policy Response on Health Care Utilization: Evidence from County-Level Medical Claims and Cellphone Data. National Bureau of Economic Research; 2020. doi:10.3386/w28131

10. Wrigley-Field E, Garcia S, Leider JP, Robertson C, Wurtz R. Racial Disparities in COVID-19 and Excess Mortality in Minnesota. Socius. 2020;6. doi:10.1177/2378023120980918

11. NCHS. AH Monthly Provisional Counts of Deaths for Select Causes of Death by Sex, Age, and Race and Hispanic Origin | Data | Centers for Disease Control and Prevention. Published 2021. Accessed August 11, 2021. https://data.cdc.gov/NCHS/AH-Monthly-Provisional-Counts-of-Deaths-for-Select/65mz-jvh5

12. Glei DA. The Us Midlife Mortality Crisis Continues: Increased Death Rates from Causes Other Than Covid-19 During 2020.; 2021:2021.05.17.21257241. doi:10.1101/2021.05.17.21257241

13. Ahmad FB, Anderson RN. The Leading Causes of Death in the US for 2020. JAMA. 2021;325(18):1829–1830. doi:10.1001/jama.2021.5469

14. US Census Bureau. National Population by Characteristics: 2010-2019. The United States Census Bureau. Published 2020. Accessed August 11, 2021. https://www.census.gov/data/tables/time-series/demo/popest/2010s-national-detail.html

15. Rossen LM. Excess Deaths Associated with COVID-19, by Age and Race and Ethnicity — United States, January 26–October 3, 2020. MMWR Morb Mortal Wkly Rep. 2020;69. doi:10.15585/mmwr.mm6942e2

16. Shiels MS, Almeida JS, García-Closas M, Albert PS, Freedman ND, Berrington de González A. Impact of Population Growth and Aging on Estimates of Excess U.S. Deaths During the COVID-19 Pandemic, March to August 2020. Ann Intern Med. 2021;174(4):437–443. doi:10.7326/M20-7385

17. Weinberger DM, Chen J, Cohen T, et al. Estimation of Excess Deaths Associated With the COVID-19 Pandemic in the United States, March to May 2020. JAMA Intern Med. 2020;180(10):1336–1344. doi:10.1001/jamainternmed.2020.3391

18. Woolf SH, Chapman DA, Sabo RT, Zimmerman EB. Excess Deaths From COVID-19 and Other Causes in the US, March 1, 2020, to January 2, 2021. JAMA. 2021;325(17):1786–1789. doi:10.1001/jama.2021.5199

19. NCHS. Underlying Cause of Death by Single Race 2018-2019. Published 2020. Accessed August 31, 2021. https://wonder.cdc.gov/wonder/help/ucd-expanded.html

20. Riley AR, Chen Y-H, Matthay EC, et al. Excess death among Latino people in California during the COVID-19 pandemic. MedRxiv Prepr Serv Health Sci. Published online January 25, 2021:2020.12.18.20248434. doi:10.1101/2020.12.18.20248434

21. Rodriguez-Diaz CE, Guilamo-Ramos V, Mena L, et al. Risk for COVID-19 infection and death among Latinos in the United States: examining heterogeneity in transmission dynamics. Ann Epidemiol. 2020;52:46-53.e2. doi:10.1016/j.annepidem.2020.07.007

22. Simon P, Ho A, Shah MD, Shetgiri R. Trends in Mortality From COVID-19 and Other Leading Causes of Death Among Latino vs White Individuals in Los Angeles County, 2011-2020. JAMA. Published online July 19, 2021. doi:10.1001/jama.2021.11945

23. Rubin-Miller L, Alban C, Sep 16 SSP, 2020. COVID-19 Racial Disparities in Testing, Infection, Hospitalization, and Death: Analysis of Epic Patient Data. KFF. Published September 16, 2020. Accessed August 12, 2021. https://www.kff.org/coronavirus-covid-19/issue-brief/covid-19-racial-disparities-testing-infection-hospitalization-death-analysis-epic-patient-data/

24. Zelner J, Trangucci R, Naraharisetti R, et al. Racial disparities in COVID-19 mortality are driven by unequal infection risks. Clin Infect Dis Off Publ Infect Dis Soc Am. Published online November 21, 2020. doi:10.1093/cid/ciaa1723

25. Rossen LM. Disparities in Excess Mortality Associated with COVID-19 — United States, 2020. MMWR Morb Mortal Wkly Rep. 2021;70. doi:10.15585/mmwr.mm7033a2

26. Price-Haywood EG, Burton J, Fort D, Seoane L. Hospitalization and Mortality among Black Patients and White Patients with Covid-19 | NEJM. N Engl J Med. 2020;382:2534–2543.

27. Kim C, Hales CM. Race and Hispanic-Origin Disparities in Underlying Medical Conditions Associated With Severe COVID-19 Illness: U.S. Adults, 2015–2018.; 2021:6.

28. Friedman J, Mann NC, Hansen H, et al. Racial/Ethnic, Social, and Geographic Trends in Overdose-Associated Cardiac Arrests Observed by US Emergency Medical Services During the COVID-19 Pandemic. JAMA Psychiatry. 2021;78(8):886–895. doi:10.1001/jamapsychiatry.2021.0967

29. Pattani A. Pandemic Unveils Growing Suicide Crisis for Communities of Color. Kaiser Health News. Published August 23, 2021. Accessed August 24, 2021. https://khn.org/news/article/pandemic-unveils-growing-suicide-crisis-for-communities-of-color/

30. Shiels MS, Tatalovich Z, Chen Y, et al. Trends in Mortality From Drug Poisonings, Suicide, and Alcohol-Induced Deaths in the United States From 2000 to 2017. JAMA Netw Open. 2020;3(9):e2016217–e2016217. doi:10.1001/jamanetworkopen.2020.16217

31. Elo IT, Hendi AS, Ho JY, Vierboom YC, Preston SH. Trends in Non-Hispanic White Mortality in the United States by Metropolitan-Nonmetropolitan Status and Region, 1990– 2016. Popul Dev Rev. 2019;45(3):549–583. doi:10.1111/padr.12249

32. Stokes AC, Lundberg DJ, Hempstead K, Bor J, Preston SH. COVID-19 and excess mortality in the United States: A county-level analysis. Published Forthcoming. Accessed July 28, 2021. https://journals.plos.org/plosmedicine/article?id=10.1371/journal.pmed.1003571

33. Pathak EB, Garcia RB, Menard JM, Salemi JL. Out-of-Hospital COVID-19 Deaths: Consequences for Quality of Medical Care and Accuracy of Cause of Death Coding. Am J Public Health. 2021;111(S2):S101-S106. doi:10.2105/AJPH.2021.306428

34. Stokes AC, Lundberg DJ, Bor J, Bibbins-Domingo K. Excess Deaths During the COVID-19 Pandemic: Implications for US Death Investigation Systems. Am J Public Health. 2021;111(S2):S53–S54. doi:10.2105/AJPH.2021.306331

35. NCHS. Guidance for Certifying Deaths Due to Coronavirus Disease 2019. U.S. Department of Health and Human Services, Centers for Disease Control and Prevention; 2020. Accessed August 15, 2021. https://asprtracie.hhs.gov/technical-resources/resource/8174/guidance-for-certifying-deaths-due-to-coronavirus-disease-2019-covid-19

